# Evaluation of triage checklist for mild COVID-19 outpatients in predicting subsequent emergency department visits and hospitalization during isolation period

**DOI:** 10.1101/2022.05.23.22275444

**Authors:** Yasuhiro Tanaka, Kazuko Yamamoto, Shimpei Morimoto, Takeshi Nabeshima, Kayoko Matsushima, Hiroshi Ishimoto, Nobuyuki Ashizawa, Tatsuro Hirayama, Kazuaki Takeda, Hiroshi Gyotoku, Naoki Iwanaga, Shinnosuke Takemoto, Susumu Fukahori, Takahiro Takazono, Hiroyuki Yamaguchi, Takashi Kido, Noriho Sakamoto, Naoki Hosogaya, Shogo Akabame, Takashi Sugimoto, Hirotomo Yamanashi, Kosuke Matsui, Mai Izumida, Ayumi Fujita, Masato Tashiro, Takeshi Tanaka, Koya Ariyoshi, Akitsugu Furumoto, Kouichi Morita, Koichi Izumikawa, Katsunori Yanagihara, Hiroshi Mukae

**Affiliations:** Department of Respiratory Medicine, Nagasaki University Graduate School of Biomedical Sciences, Nagasaki City, Nagasaki, 852-8501, Japan; Department of Respiratory Medicine, Nagasaki University Hospital, Nagasaki City, Nagasaki, 852-8501, Japan; Clinical Research Center, Nagasaki University Hospital, Nagasaki City, Nagasaki, 852-8102, Japan; Department of Virology, Institute of Tropical Medicine, Nagasaki University, Nagasaki City, Nagasaki, 852-8523, Japan; Medical Education Development Center, Nagasaki University Hospital, Nagasaki City, Nagasaki, 852-8501, Japan; Infection Control and Education Center, Nagasaki University Hospital, Nagasaki City, Nagasaki, 852-8501, Japan; Department of Infectious Diseases, Nagasaki University Graduate School of Biomedical Sciences, Nagasaki City, Nagasaki, 852-8501, Japan; Clinical Oncology Center, Nagasaki University Hospital, Nagasaki City, Nagasaki, 852-8501, Japan; Department of General Medicine, Nagasaki University Hospital, Nagasaki City, Nagasaki, 852-8501, Japan; Department of Infectious Diseases, Nagasaki University Hospital, Nagasaki City, Nagasaki, 852-8523, Japan; Infectious Diseases Experts Training Center, Nagasaki University Hospital, Nagasaki City, Nagasaki, 852-8501, Japan; Department of Laboratory Medicine, Nagasaki University Hospital, Nagasaki City, Nagasaki, 852-8501, Japan

**Keywords:** COVID-19, outpatient, triage checklist, emergency department visit, hospitalization

## Abstract

**Background and objective:** Limited evidence exists regarding the outcomes of patients with coronavirus disease 2019 (COVID-19) who are not hospitalized. This study aimed to assess the outcomes for mild COVID-19 patients in terms of emergency department (ED) visits and hospital admission given initial outpatient triage evaluation and to identify the triage factors affecting these outcomes.

**Methods:** This retrospective cohort study investigated adult COVID-19 Japanese patients who were triaged at Nagasaki University Hospital between April 1, 2021, and May 31, 2021. A triage checklist with 30 factors was used to identify patients requiring hospitalization. Patients recommended for isolation were followed up for later ED visit or hospital admission.

**Results:** Overall, 338 COVID-19 patients (mean age, 44.7; 45% women) visited the clinic at an average of 5.4 days after symptom onset. Thirty-six patients (10.6%) were hospitalized from triage, and the rest were recommended for isolation. Seventy-two non-hospitalized patients (23.8%) visited ED during their isolation period, and 30 (9.9%) were hospitalized after ED evaluation. The mean duration to ED visit and hospitalization after symptom onset were 8.8 and 9.7 days, respectively. Checklist factors associated with hospitalization during the isolation period were age > 50 years, obesity with BMI > 25, underlying hypertension, tachycardia with HR > 100/min or blood pressure >135 mmHg at triage, and >□3-day delay in hospital visit after symptom onset.

**Conclusion:** Clinicians should be wary of COVID-19 patients with above risk factors and prompt them to seek follow-up assessment by a medical professional.

**SUMMARY AT A GLANCE:** Overall, 338 patients with mild COVID-19 were retrospectively followed up. Factors such as age >□50 years, BMI□> □25, underlying hypertension, high blood pressure and tachycardia at triage, and delayed visit after symptom onset were associated with emergency department visit and hospitalization during the isolation period.

## INTRODUCTION

In January 2020, coronavirus disease 2019 (COVID-19)—caused by the severe acute respiratory syndrome coronavirus-2 (SARS-CoV-2)—first appeared in Japan, and it has caused five pandemic waves counting to the end of 2021. Overall, 2.93 million polymerase chain reaction (PCR)-positive cases have been confirmed, and 18,393 deaths have been reported by the Ministry of Health, Labour and Welfare in Japan as of December 31, 2021.^1^ During the COVID-19 pandemic, emergency departments (EDs) were involved in the crisis, managing cases of severe respiratory failure as well as a large number of patients with mild symptoms.^2^ This caused overcrowding in many EDs and unavailability of hospital beds across the nation.^3^ An early triage system is critical for identifying patients with high risk of severe COVID-19 who will require hospitalization.^4^ To date, many risk factors have been identified that contribute to the severity of COVID-19, such as advanced age, obesity, and diabetes mellitus.^5^ However, there is minimal evidence regarding the outcomes of patients with mild COVID-19 who are not hospitalized and under home or facility isolation. Factors associated with later exacerbation of symptoms, leading to an ED visit and admission of these patients, have not been clarified. Although more than 80% of patients with COVID-19 have mild disease,^6^ this percentage has further increased following the introduction of SARS-CoV-2 vaccinations.^7^ Investigation of patients with mild disease is important to avoid disruption of EDs and the society at large. The present study aimed to assess the outcomes of mild COVID-19 patients in terms of subsequent ED visits and hospitalization following initial triage and identify the factors leading to these outcomes. Its novelty is contributing an evaluation of the potential benefits of a triage checklist in avoiding treatment delay and subsequent hospitalization of patients with mild COVID-19.

## METHODS

### Study population

This was a retrospective, observational cohort study of patients aged ≥□20 years who visited the COVID-19 triage outpatient clinic at Nagasaki University Hospital (NUH) between April 1, 2021, and May 31, 2021, during Japan’s fourth wave of the COVID-19 pandemic. Since the pandemic, a COVID-19 triage outpatient clinic has been operational at NUH for patients with mild illnesses. SARS-CoV-2 PCR-positive patients detected in Nagasaki City are listed by the Nagasaki City Public Health Center and reported to the Nagasaki University Hospital Infection Control and Education Center (NICE), which then assigns these patients to hospitals for initial assessment. NUH caters to the largest number of COVID-19 patients in Nagasaki City. Such patients arriving at NUH for first assessment are examined by a doctor in a triage room or tent and interviewed using a COVID-19 checklist while a nurse records their vital signs. Patients who directly visited or were transported to the emergency room for hospital admission without outpatient triage were excluded from this analysis.

### Factors in the triage checklists

Checklist factors included age, sex, date of symptom onset, height, weight, body mass index (BMI), vital signs (body temperature [BT], respiratory rate [RR], SpO_2_, pulse rate [PR], systolic blood pressure [sBP], diastolic blood pressure [dBP]), underlying diseases (hypertension, diabetes, cardiovascular diseases, chronic lung diseases, chronic liver diseases, chronic renal diseases, malignancies, history of organ transplantation, human immunodeficiency virus [HIV] infection, use of oral steroids/immunosuppressive drugs, and pregnancy), and symptoms that may be considered for hospitalization (fatigue with difficulty moving, dyspnea, chest pain, poor dietary intake, and severe diarrhea with ≥□6 bowel movements per day). Indication for hospitalization was determined by a doctor based on the checklist. Hospitalization was recommended if the patient’s SpO_2_ was ≤□95%, RR was ≥□22/min, sBP was ≤ □90 mmHg, or if severe dyspnea was present.

### SARS-CoV-2 genetic background in the study period at Nagasaki prefecture

The epidemic since March 2021 in Japan is called the “fourth wave.” To clarify the genotypic changes in the fourth wave, we constructed a phylogenetic tree of SARS-CoV-2 isolated in Nagasaki Prefecture. Eighteen hundred ten (1810) records of SARS-CoV-2 genome sequences from Nagasaki Prefecture were available on GISAID^8^ as of 2022/03/24. The tree was generated by Nextstrain.^9.10^ The third wave in Japan lasted until June 2020. COVID-19 was not endemic in Nagasaki Prefecture after the third wave until the end of March 2021. The phylogenetic tree (Supplementary Fig. 1) suggested that a clade 20B/R.1 had remained in Nagasaki Prefecture during the low endemic period. The “fourth wave” is mainly caused by the clade 20I/B.1.1.7/Alpha. This clade was first detected in Japan in December 2020 and introduced into Nagasaki Prefecture until late March 2021. The tree suggests that at least four independent sub-clades of 20I/B.1.1.7/Alpha were introduced into Nagasaki Prefecture around the end of March 2021. And the old clade, 20B/R.1, remained in Nagasaki Prefecture until May 2021. Nagasaki City is located in the southern part of Nagasaki Prefecture. The diversity of genomic RNA sequences suggested that 20I/B.1.1.7/Alpha spread rapidly in Nagasaki Prefecture beginning in early April 2021. However, many of the GISAID records do not include geographical information in Nagasaki Prefecture. Therefore, it is possible that some of the cases investigated in this study may have been caused by the older 20B/R.1.

### End point

The outcomes of the present study were 1) ED visit during isolation at home or facilities, and 2) hospitalization following ED visit.

### Ethics

This study was approved by the Ethical Review Committee of NUH (permission number 21101927), and the study was conducted in accordance with the Declaration of Helsinki. The research was conducted in an opt-out format, and the information was disclosed at the Clinical Research Center, NUH.

### Analysis

All variables in the COVID-19 triage checklist were included in the analysis. Events were designated as “ED visit during isolation” or “hospitalization following ED visit,” and the association of a variable with each of these events was described as the odds ratio or the mean difference. The p-value for the null hypothesis for each of the associations, derived from either Fisher’s exact test or Wilcoxon rank sum test, was utilized as a signal. We stated “significantly” associated when the obtained *p*-value was lower than 0.05.

## RESULTS

### Patients’ characteristics at outpatient triage

The period of the study was applied to the fourth wave of COVID-19 pandemic in Japan, which was due to mainly the clade 20I/B.1.1.7/Alpha and partly the old clade, 20B/R.1 (Figure S1). Overall, 338 SARS-CoV-2-positive patients visited the NUH outpatient triage clinic, and their characteristics are listed in Table 1. All patients were of Japanese ethnicity with a mean age of 44.7 ± 15.2 years, and 45% of them were women. Outpatient triage visits occurred at a mean of 5.4 days after symptom onset. The mean values and standard deviation for BMI, BT, RR, SpO_2_, PR, and sBP/dBP at triage were, 23.6 ± 4.4 kg/m^2^, 37.0 ± 0.7□, 17.9 ± 5.6/min, 96.8 ± 3.0%, 87.0 ± 14.4/min, and 129 ± 20.7/76 ± 13.6 mmHg), respectively. Overall, 71 patients (21.0%) had underlying diseases. The most frequent comorbidity was hypertension (11%), followed by chronic lung diseases (4.4%), cardiovascular diseases (3.8%), and diabetes (3.6%). Of all the patients, 2.1% were pregnant. None of the patients had a history of organ transplant or HIV infection. Symptoms that potentially necessitated hospitalization were found in 111 patients (32.8%). Poor dietary intake (22.0%) was the most frequent symptom calling for hospitalization, followed by dyspnea (8.3%) and chest pain (6.5%). Of the 338 patients evaluated, 36 patients (10.6%) were hospitalized at the initial triage, while the remaining 302 patients were isolated in facilities or at home (Figure 1). To assess the factors in the checklist relating to patients’ admissions, the group of patients hospitalized at initial triage was compared with those who underwent facility- or home-isolation. The factors that were likely associated (*p* <□.1) with hospitalization at triage are shown in Table 2. Patients hospitalized at triage were likely to consult at a later time (6.2 days) after symptom onset compared to those who underwent isolation (5.3 days). The former group of patients were also significantly older (mean age, 58.2 years; first quartile, 50.8 years) and had a higher BMI (mean, 26.1 kg/m^2^) than the non-hospitalized group. Moreover, the following clinical factors were associated with the hospitalization at triage: high fever, tachypnea, oxygen desaturation, and tachycardia. BT was the only factor that did not overlap between the hospitalized (first quartile, 37.6) and non-hospitalized groups (third quartile, 37.3). Comorbidities such as hypertension, diabetes, cardiovascular diseases, or liver diseases were associated with hospitalization at triage. Patients hospitalized at triage had significant symptoms of severe fatigue, dyspnea, or poor dietary intake. The outcomes of patients hospitalized at initial triage were as follows: three patients (8.3%) did not progress from mild disease; 11 patients (30.6%) progressed to non-oxygen requiring pneumonia; 21 patients (58.3%) progressed to moderate pneumonia requiring oxygen supplementation; and one patient (2.8%) died from severe disease (Figure 1).

**Table 1.** Characteristics of COVID-19 patients at outpatient triage.

| factors in the triage check sheet | mean | Q1<br>(25%) | Q2<br>(50%) | Q3<br>(75%) |
| --- | --- | --- | --- | --- |
| Days post symptom onset | 5.4 | 3.8 | 5.0 | 7.0 |
| Age | 44.7 | 32 | 44 | 56 |
| Female sex | 152 (45.0%) |  |  |  |
| BMI (kg/m<sup>2</sup>) | 23.6 | 20.8 | 22.9 | 25.8 |
| BT (□) | 37.0 | 36.5 | 36.9 | 37.4 |
| RR (/min) | 17.9 | 16.0 | 16.0 | 20.0 |
| SpO<sub>2</sub> (%) | 96.8 | 96.0 | 97.0 | 98.0 |
| PR (/min) | 87.0 | 78.0 | 86.0 | 98.0 |
| sBP (mmHg) | 129.0 | 114.0 | 128.0 | 142.0 |
| dBp (mmHg) | 76.4 | 67.0 | 77.0 | 85.0 |
| factors in the triage check sheet | n | % |  |  |
| Underlying diseases | 71 | 21.0 |  |  |
| Hypertension | 37 | 11.0 |  |  |
| Diabetes mellitus | 12 | 3.6 |  |  |
| Cardiovascular diseases | 13 | 3.8 |  |  |
| Chronic lung diseases | 15 | 4.4 |  |  |
| Chronic liver diseases | 3 | 0.9 |  |  |
| Chronic renal diseases | 2 | 0.6 |  |  |
| Malignancy | 5 | 1.4 |  |  |
| Organ transplant | 0 | 0 |  |  |
| HIV infection | 0 | 0 |  |  |
| Other immunosuppressed status | 5 | 1.4 |  |  |
| Pregnancy | 7 | 2.1 |  |  |
| factors in the triage check sheet | n | % |  |  |
| Symptoms with consideration for hospitalization | 111 | 32.8 |  |  |
| fatigue with difficulty moving | 14 | 4.1 |  |  |
| dyspnea | 28 | 8.3 |  |  |
| chest pain | 22 | 6.5 |  |  |
| poor dietary intake | 76 | 22.0 |  |  |
| severe diarrhea (≥ 6 times/ day) | 6 | 1.8 |  |  |

Abbreviations; ED, emergency department; BMI, body mass index; BT, body temperature; RR, respiratory rate; PR, pulse rate; sBP, systolic blood pressure; dBp, diastolic blood pressure; HIV, human immunodeficiency virus; Q1, first quartile; Q2, second quartile; Q3, third quartile.

**Table 2.** Checklist factors and hospitalization at outpatient triage.

|  | mean | Q1<br>(25%) | Q2<br>(50%) | Q3<br>(75%) | *P* value |
| --- | --- | --- | --- | --- | --- |
| Days post symptom onset |  |  |  |  | 0.061 |
| Hospitalized | 6.2 | 4.5 | 5.0 | 7.5 |  |
| Isolation on facilities or home | 5.3 | 3.0 | 5.0 | 7.0 |  |
| Age |  |  |  |  | <0.001 |
| Hospitalized | 58.2 | 50.8 | 58.5 | 67.0 |  |
| Isolation on facilities or home | 43.1 | 31.0 | 41.0 | 53.0 |  |
| BMI (kg/m<sup>2</sup>) |  |  |  |  | 0.002 |
| Hospitalized | 26.1 | 22.8 | 26.1 | 28.2 |  |
| Isolation on facilities or home | 23.4 | 20.5 | 22.7 | 25.4 |  |
| BT (°C) |  |  |  |  | <0.001 |
| Hospitalized | 37.7 | 37.6 | 38.5 | 40.0 |  |
| Isolation on facilities or home | 37.0 | 36.5 | 36.9 | 37.3 |  |
| RR (/min) |  |  |  |  | 0.002 |
| Hospitalized | 20.8 | 16.0 | 18.0 | 25.5 |  |
| Isolation on facilities or home | 17.6 | 16.0 | 16.0 | 20.0 |  |
| SpO<sub>2</sub> (%) |  |  |  |  | <0.001 |
| Hospitalized | 93.4 | 94.0 | 95.0 | 97.0 |  |
| Isolation on facilities or home | 97.2 | 97.0 | 97.0 | 98.0 |  |
| PR (/min) |  |  |  |  | 0.004 |
| Hospitalized | 94.4 | 84.8 | 94.0 | 105.3 |  |
| Isolation on facilities or home | 86.2 | 78.0 | 85.0 | 96.0 |  |
|  |  | Hospitalized<br>(n=36) | Isolation on<br>facilities or<br>home<br>(n=302) | Odds ratio | *P* value |
| Hypertension |  | 11 (30.6) | 26 (8.6) | 1.85-11.17 | 0.001 |
| Cardiovascular diseases |  | 4 (11.1) | 9 (3.0) | 0.86-15.53 | 0.039 |
| Diabetes mellitus |  | 4 (11.1) | 8 (2.6) | 0.95-18.20 | 0.029 |
| Chronic liver diseases |  | 2 (5.6) | 1 (0.3) | 0.88-1039.73 | 0.031 |
| Severe fatigue with hard to<br>move |  | 5 (13.9) | 9 (3.0) | 1.29-18.64 | 0.010 |
| Malignancy |  | 2 (5.6) | 3 (1.0) | 0.47-52.58 | 0.090 |
| Immunosuppressed status |  | 2 (5.6) | 3 (1.0) | 0.47-52.58 | 0.090 |
| dyspnea |  | 14 (38.9) | 14 (4.6) | 5.02-33.50 | <0.001 |
| poor dietary intake |  | 14 (38.9) | 62 (20.5) | 1.09-5.36 | 0.013 |

Abbreviations; BMI, body mass index; BT, body temperature; RR, respiratory rate; PR, pulse rate; Q1, first quartile; Q2, second quartile; Q3, third quartile.

**Figure 1.**
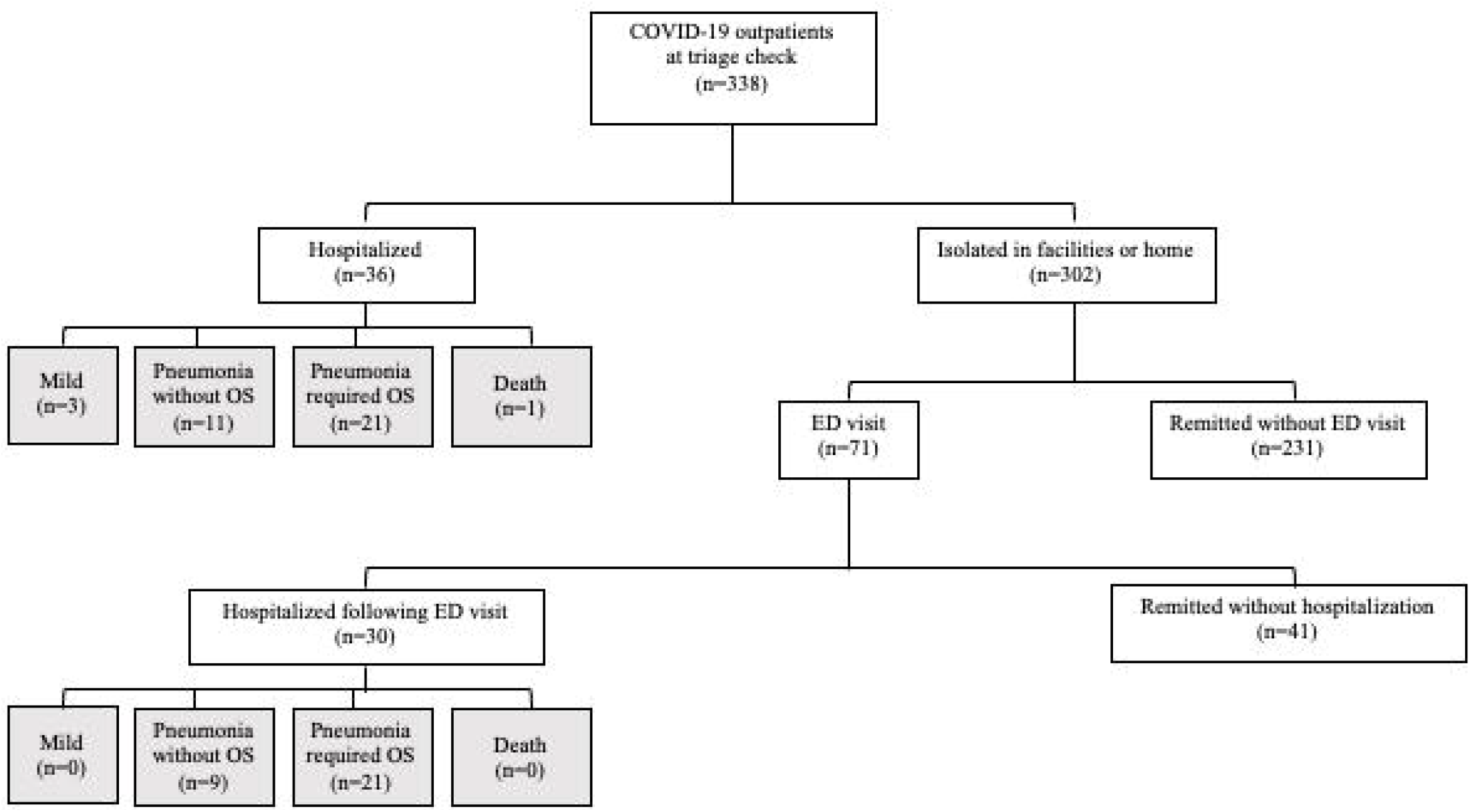
Flowsheet demonstrating the numbers of COVID-19 patients who visited the outpatient triage, who were hospitalized at triage, who visited the emergency department (ED) during isolation, and who were hospitalized at ED visit during isolation (white-filled square). Hospitalized patients were followed up and outcomes were recorded (gray-filled square). Abbreviation; OS, oxygen supplementation.

### Triage factors associated with ED visit during isolation at facilities or home

Among the 302 patients who were isolated in facilities or at home, 71 patients (23.5%) visited the ED during the isolation period (Figure 1). The mean duration from symptom onset to ED visit was 8.8 ±2.5 days. Patients who visited the ED while on isolation and those who did not visit the ED while on isolation were compared. Table 3 lists the factors that were likely associated (*p* □<□0.1) with ED visits while in facility- or home-isolation. ED visit during isolation was associated with older age (among these without ED visit, 75% patients were ≤□<u>□</u>50 years old, while among those who visited the ED, 50% were >□50 years old) and higher BMI (75% of patients without ED visit had BMI <□24.9 kg/m^2^, while 50% of the patients who visited ED had <u>>□</u>□24.4 kg/m^2^). Moreover, low grade fever (75% of patients without ED visit had BT < □37.2, while 50% of patients who visited ED had BT ≥ □37.1) and higher sBP (75% patients without ED visit had sBP ≤ □139 mmHg, while 50% of patients who visited ED had sBP ≥□134 mmHg) at triage were associated with ED visit during isolation. Lower SpO_2_ and higher PR were statistically likely associated with ED, although there were small differences in the variance. A total of 16.9% of patients had hypertension, while 12.7% reported having chest pain at triage.

**Table 3.** Checklist factors associated with ED visit during isolation on facilities or home.

|  | mean | Q1<br>(25%) | Q2<br>(50%) | Q3<br>(75%) | *P* value |
| --- | --- | --- | --- | --- | --- |
| Age |  |  |  |  | <0.001 |
| ED visit | 49.6 | 35.5 | 50.0 | 61.5 |  |
| Remission without ED visit | 41.1 | 29.0 | 40.0 | 50.0 |  |
| BMI (kg/m<sup>2</sup>) |  |  |  |  | 0.044 |
| ED visit | 24.2 | 21.6 | 24.4 | 26.2 |  |
| Remission without ED visit | 23.1 | 20.2 | 22.3 | 24.9 |  |
| BT (°C) |  |  |  |  | <0.001 |
| ED visit | 37.2 | 36.7 | 37.1 | 37.7 |  |
| Remission without ED visit | 36.9 | 36.5 | 36.8 | 37.2 |  |
| SpO<sub>2</sub> (%) |  |  |  |  | 0.042 |
| ED visit | 97.0 | 96.0 | 97.0 | 98.0 |  |
| Remission without ED visit | 97.3 | 97.0 | 97.0 | 98.0 |  |
| PR (/min) |  |  |  |  | 0.014 |
| ED visit | 89.8 | 79.0 | 87.0 | 99.5 |  |
| Remission without ED visit | 85.0 | 77.0 | 84.0 | 95.0 |  |
| sBP (mmHg) |  |  |  |  | 0.008 |
| ED visit | 134.5 | 121.0 | 134.0 | 147.0 |  |
| Remission without ED visit | 126.7 | 113.0 | 126.0 | 139.0 |  |
|  |  | ED visit<br>(n=71) | Remission<br>without ED<br>visit<br>(n=231) | Odds ratio | *P* value |
| Hypertension |  | 12 (16.9) | 14 (6.0) | 1.25-7.76 | 0.004 |
| chest pain |  | 9 (12.7) | 10 (4.3) | 1.10-9.18 | 0.021 |

Abbreviations; ED, emergency department; BMI, body mass index; BT, body temperature; PR, pulse rate; sBP, systolic blood pressure; Q1, first quartile; Q2, second quartile; Q3, third quartile.

### Triage factors associated with hospital admission at ED visit during isolation at facilities or home

Among the 302 patients who were isolated to facilities or home after triage, 30 patients (9.9%) were hospitalized after an ED visit during the isolation period (Figure 1). To assess which factors at triage distinguished patients who required hospitalization during isolation, we compared the characteristics of patients who were hospitalized at ED visit and those who were not hospitalized at ED visit. Factors that were likely associated (*p* <□.1) with hospitalization at ED during the isolation period shown in Table 4. The mean duration from COVID-19 symptom onset to hospitalization was 9.7 ±2.0 days. Patients hospitalized at ED visit were likely to present at initial triage later (5.5 days) than patients in the non-hospitalized group (4.5 days) post symptomatic. Duration from first triage to ED visit was 2.0 ±2.5 days. Patients who were hospitalized following ED visit during isolation were older (75% of patients without hospitalization at ED visit were <□52 years old while 75% of patients who were hospitalized at ED were > 49.5 years old) and had higher BMI (75% of patients without hospitalization at ED visit had BMI <□25.9 kg/m^2^, while 50% of patients who were hospitalized at ED visit had BMI >□25.3 kg/m^2^). At triage, patients who were hospitalized following ED visit during isolation had low grade fever (75% of patients without ED visit had BT <□37.4□, while 50% of patients who visited ED had >□37.5□) and higher sBP (75% of patients without ED visit had sBP <□143 mmHg, while 50% of patients who visited ED had sBP >□140 mmHg). A total of 30% of patients who were hospitalized at ED visit had underlying hypertension. The outcomes of patients hospitalized following ED visit were as follows: none maintained their mild disease (0%); 9 (30.0%) progressed to non-oxygen requiring pneumonia; 21 (70.0%) progressed to oxygen-requiring pneumonia; and none died or progressed to severe disease requiring intensive care unit admission (Figure 1).

**Table 4.** Checklist factors associated with hospitalization following ED visit during isolation.

|  | mean | Q1<br>(25%) | Q2<br>(50%) | Q3<br>(75%) | *P* value |
| --- | --- | --- | --- | --- | --- |
| Days post symptom onset |  |  |  |  | 0.028 |
| Hospitalized at ED | 5.5 | 4.0 | 5.0 | 6.0 |  |
| Non-hospitalized at ED | 4.5 | 3.0 | 4.0 | 6.0 |  |
| Age |  |  |  |  | <0.001 |
| Hospitalized at ED | 57.9 | 49.5 | 58.5 | 69.5 |  |
| Non-hospitalized at ED | 43.5 | 32.0 | 39.0 | 52.0 |  |
| BMI (kg/m<sup>2</sup>) |  |  |  |  | 0.092 |
| Hospitalized at ED | 25.0 | 22.3 | 25.3 | 26.8 |  |
| Non-hospitalized at ED | 23.5 | 21.3 | 23.4 | 25.9 |  |
| Days post symptom onset |  |  |  |  | 0.028 |
| Hospitalized at ED | 5.5 | 4.0 | 5.0 | 6.0 |  |
| Non-hospitalized at ED | 4.5 | 3.0 | 4.0 | 6.0 |  |
| BT (°C) |  |  |  |  | 0.008 |
| Hospitalized at ED | 37.4 | 37.0 | 37.5 | 37.9 |  |
| Non-hospitalized at ED | 37.0 | 36.6 | 37.0 | 37.4 |  |
| SpO<sub>2</sub> (%) |  |  |  |  | 0.002 |
| Hospitalized at ED | 96.4 | 96.0 | 96.5 | 97.0 |  |
| Non-hospitalized at ED | 97.4 | 97.0 | 97.0 | 98.0 |  |
| PR (/min) |  |  |  |  | 0.055 |
| Hospitalized at ED | 93.6 | 82.5 | 94.5 | 103.0 |  |
| Non-hospitalized at ED | 87.0 | 78.0 | 84.0 | 97.0 |  |
| sBP (mmHg) |  |  |  |  | 0.064 |
| Hospitalized at ED | 140.4 | 125.5 | 140.0 | 148.8 |  |
| Non-hospitalized at ED | 130.2 | 113.0 | 127.5 | 143.0 |  |
|  |  | Hospitalized<br>at ED<br>(n=30) | Non-hospitalized<br>at ED<br>(n=41) | Odds ratio | *P* value |
| Female Sex |  | 9 (30.0) | 21 (51.2) | 0.82-7.55 | 0.074 |
| Hypertension |  | 9 (30.0) | 3 (7.3) | 1.15-33.68 | 0.012 |

Abbreviations; ED, emergency department; BMI, body mass index; BT, body temperature; PR, pulse rate; sBP, systolic blood pressure; Q1, first quartile; Q2, second quartile; Q3, third quartile.

A heatmap for variables comparing those who were hospitalized following ED visit (n□=□30) to those were not hospitalized (n□=□272) during isolation is shown in Figure 2. The OR of hospitalization and the p-value (null hypothesis: OR□=□1) were obtained for each variable. The continuous variables were dichotomized at the cutoffs stated by Youden index. Age >□50 years (OR□=□6.8), obesity with BMI > 25 kg/m^2^ (OR□=□3.2), underlying hypertension (OR□=□6.4), tachycardia with PR□>□100/min (OR□=□2.7), high blood pressure (sBP□>□135 mmHg) at triage (OR□=□3.5), and delay of presence to hospital >□3 days after symptom onset (OR□=□3.5) were associated with hospitalization during isolation period. To consider confounding and other variables related to delay of triage visit (“duration” in the Figure 2), the distribution of the delay was explored using the relative rank of the numbers of days. The result showed that patients aged 65 years or older and of male sex had a tendency for later triage visit (median, 1.5 days) compared to those younger than 65 years or of female sex (median, 1.0 days) (Supplemental Figure 2A and 2B).

**Figure 2.**
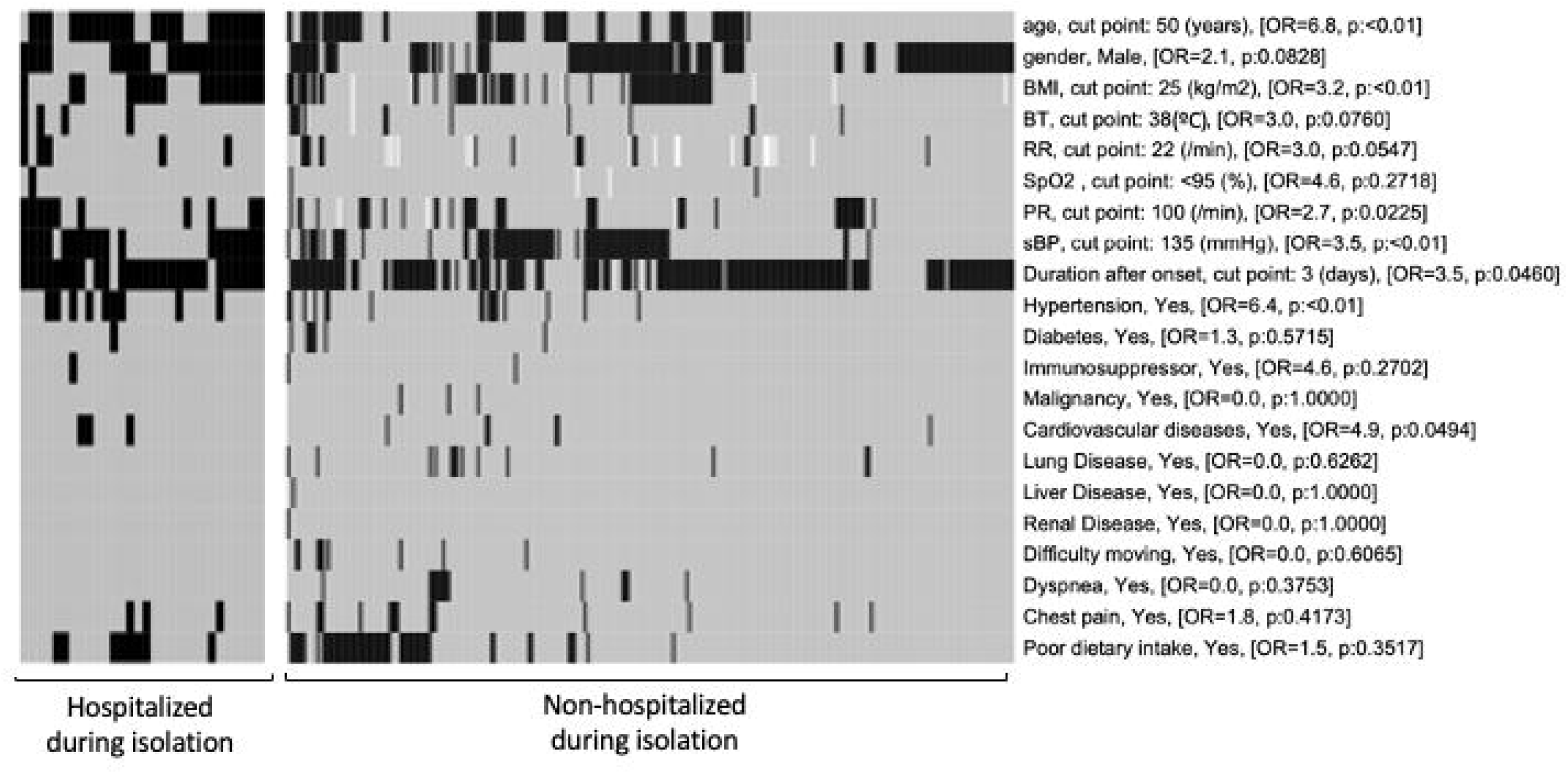
Heatmap on each variable in the COVID-19 outpatient triage. Hospitalized patients at ED visit (n□=□30) and patients without hospitalization during isolation (n□=□272) were grouped. Black-filled patients represent values over than cutoff values (for continuous variable) or presence of underlying diseases or symptoms. Gray-filled patients represent under than cutoff values (for continuous variable) or absence of underlying diseases or symptoms. White-filled patients represent missing data. The cutoff value was defined using Youden index. In the brackets, odds ratio (OR) and p values (Fisher exact test) for the association between the variable (≤□median, >□median) and occurrence of “Hospitalized at ED visit”. Abbreviations; BMI, body mass index; BT, body temperature; RR, respiratory rate; PR, pulse rate; sBP, systolic blood pressure.

## DISCUSSION

In this retrospective cohort study of 338 SARS-CoV-2-positive patients, we provide evidence to state that relatively young patients (age < 53 years) and those without hypertension at triage can self-manage their conditions without hospitalization. Our sample had a higher rate of ED visits than the 14% rate reported by a previous study, which followed patients with coronavirus-like symptoms during COVID-19 pandemic.^11^ Our study results show that >□10% of the patients were hospitalized at first triage visit and that another 10% were hospitalized following an ED visit during isolation; these proportions were also similar when compared to previous reports.^12,13^ Although all patients had only mild disease at initial triage visit, the condition of those who were hospitalized progressed to pneumonia, with 60–70% of them requiring oxygen therapy and one patient dying during hospitalization. High prevalence of disease progression and hospitalization in our cohort may have been caused by the alpha variant, which was reported to be associated with increased hospitalization risk.^14^ The clinical course of disease progression to ED visit or hospitalization in our cohort was similar to that reported in earlier publications, which described progression to maximal symptom severity at 8–10 days after symptom onset.^5^ The Ministry of Health, Labour and Welfare of Japan has specified that at least 10 days of isolation is required after the onset of COVD-19 symptoms to prevent transmission,^16^ which is a reasonable time for patients to observe their symptoms for possible disease progression. In our study, the mean interval from first triage to ED return was 2 days, which is much shorter than the 5 days reported previously.^11^ We attribute this result to the dense monitoring system of Nagasaki City Public Health Center or isolation facilities. Our results suggested that later triage visit after symptom onset was associated with ED visit or hospitalization during the isolation period. Similar to previous reports,^17,18^ a delay in hospital visit was seen in patients who were >□65 years of age or male. This finding may reflect the difference in health seeking behavior across sex and age groups and/or the non-recognition of developing symptoms in the elderly.^19^ Our study indicated that COVID-19 patients aged□50 years, those with obesity per BMI >□25, those having underlying hypertension, those presenting to the hospital >□3 days after symptom onset, and those presenting with tachycardia (PR >□100/min)^2^ or high blood pressure (sBP >□135 mmHg) at triage significantly required admission during isolation. COVID-19 outpatients with the abovementioned features at first triage should be better followed up remotely^20^ or by phone^21^ to avoid missing their proper hospital visit.

The study has several limitations. First, this was a retrospective study and, therefore, has a potential for biases from incomplete clinician documentation. Second, follow-up information was not available after patients were discharged, which may have led to underestimated rates of hospitalization or mortality. Third, members of the team reviewing the ED revisits were not blinded, which may have introduced assessment bias. Fourth, since different variants of coronavirus have different clinical characteristics, important factors at the triage may be different during other pandemic waves. Lastly, the generalizability of the current study findings to other settings is limited by single-center design. Because of site-specific characteristics, it is possible that other ED sites with differences in availability of transportation and access to professional medical support may have different rates of outcomes than our own patient cohort.

In summary, approximately 80% of patients with mild COVID-19 disease can be safely isolated at home in a facility. Ten percent of patients will experience progression of symptoms in the ensuing week that will require hospitalization for treatment—typically at 10 days of symptomatic illness. Clinicians should inform patients, especially those aged >□50 years, or those that are obese with BMI > 25, or those with underlying hypertension, or those presenting to the hospital > 3 days after symptom onset and presenting tachycardia (PR > 100/min) or high blood pressure (sBP > 135 mmHg) at triage, that they may experience worsening of symptoms after the first visit and may eventually require hospitalization. Such patients should be advised to seek follow-up assessment by a medical professional. Further prospective studies in the upcoming new COVID-19 pandemic waves will be recommended to clarify checklist factors and cutoff values that affect patients’ outcomes.

## Data Availability

All data produced in the present work are contained in the manuscript

## ACKNOWLEDGEMENTS

We acknowledge all medical staff of the outpatient triage clinic in Nagasaki University Hospital for their care provided to COVID-19 patients.

## Author Contributions

Conceptualization: Kazuko Yamamoto;

Data curation: Yasuhiro Tanaka, Kazuko Yamamoto, and Shimpei Morimoto;

Formal analysis: Shimpei Morimoto and Takeshi Nabeshima;

Funding acquisition: Kazuko Yamamoto and Hiroshi Mukae;

Investigation: Kayoko Matsushima, Hiroshi Ishimoto, Nobuyuki Ashizawa, Tatsuro Hirayama, Kazuaki Takeda, Hiroshi Gyotoku, Naoki Iwanaga, Shinnosuke Takemoto, Naoyuki Yamaguchi, Susumu Fukahori, Takahiro Takazono, Hiroyuki Yamaguchi, Takashi Kido, Noriho Sakamoto, Naoki Hosogaya, Shogo Akabame, Takafumi Sugimoto, Hirotomo Yamanashi, Kosuke Matsui, Mai Izumida, Masato Tashiro, Takeshi Tanaka, Koya Ariyoshi, and Akitsugu Furumoto;

Methodology: Kazuko Yamamoto and Shimpei Morimoto;

Project administration: Kazuko Yamamoto and Hiroshi Mukae;

Resources: Koichi Izumikawa and Hiroshi Mukae;

Supervision: Katsunori Yanagihara and Kouichi Morita;

Validation: Kazuko Yamamoto and Shimpei Morimoto;

Writing—original draft: Yasuhiro Tanaka;

Writing—review & editing: Kazuko Yamamoto.

## FIGURE LEGEND

**Supplemental Figure 1.**
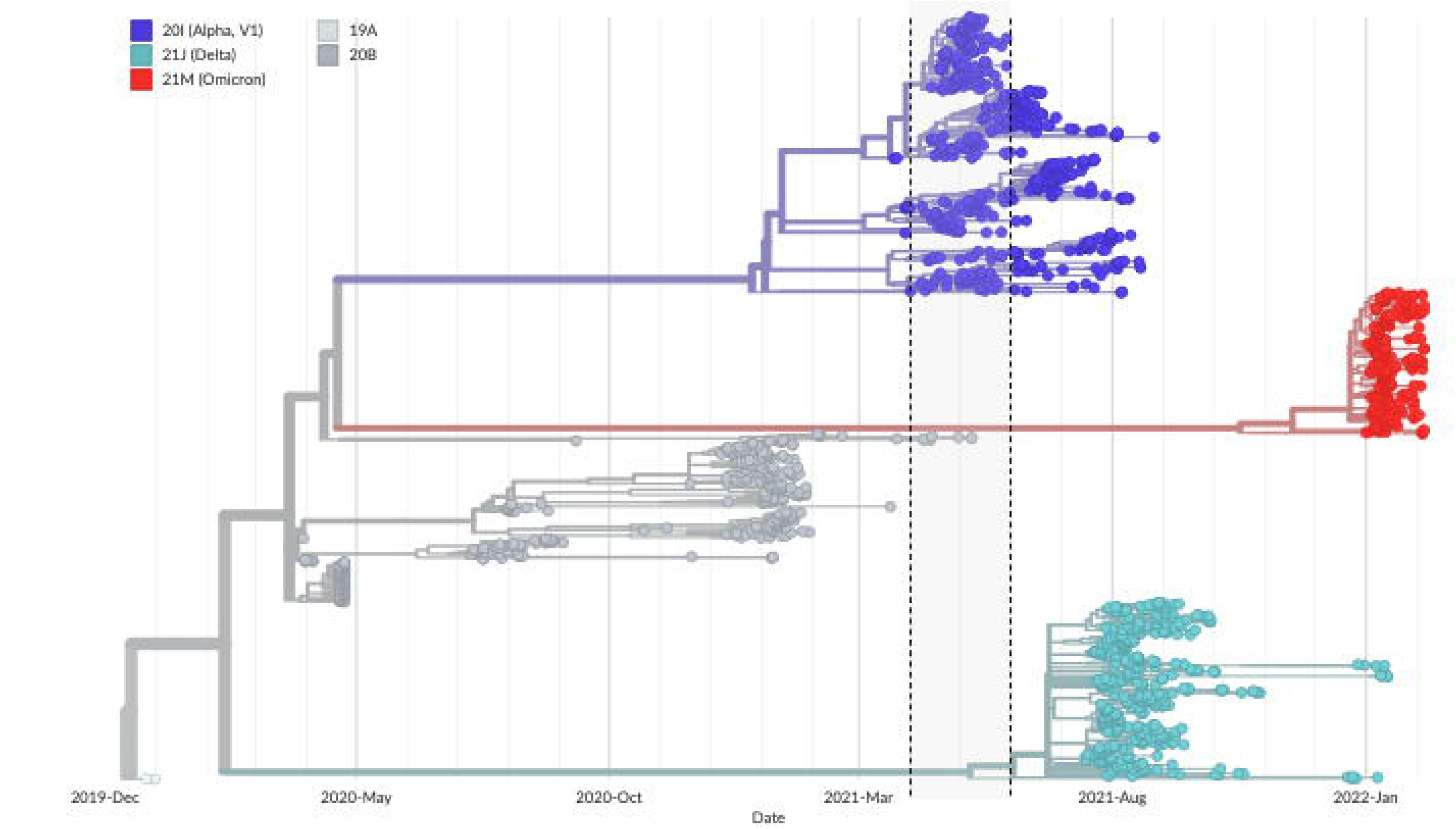
Phylogenetic tree of SARS-COV-2 which isolated in Nagasaki prefecture. Data were obtained from GISAID. The area enclosed by the dotted line is the time period of this study.

**Supplemental Figure 2.**
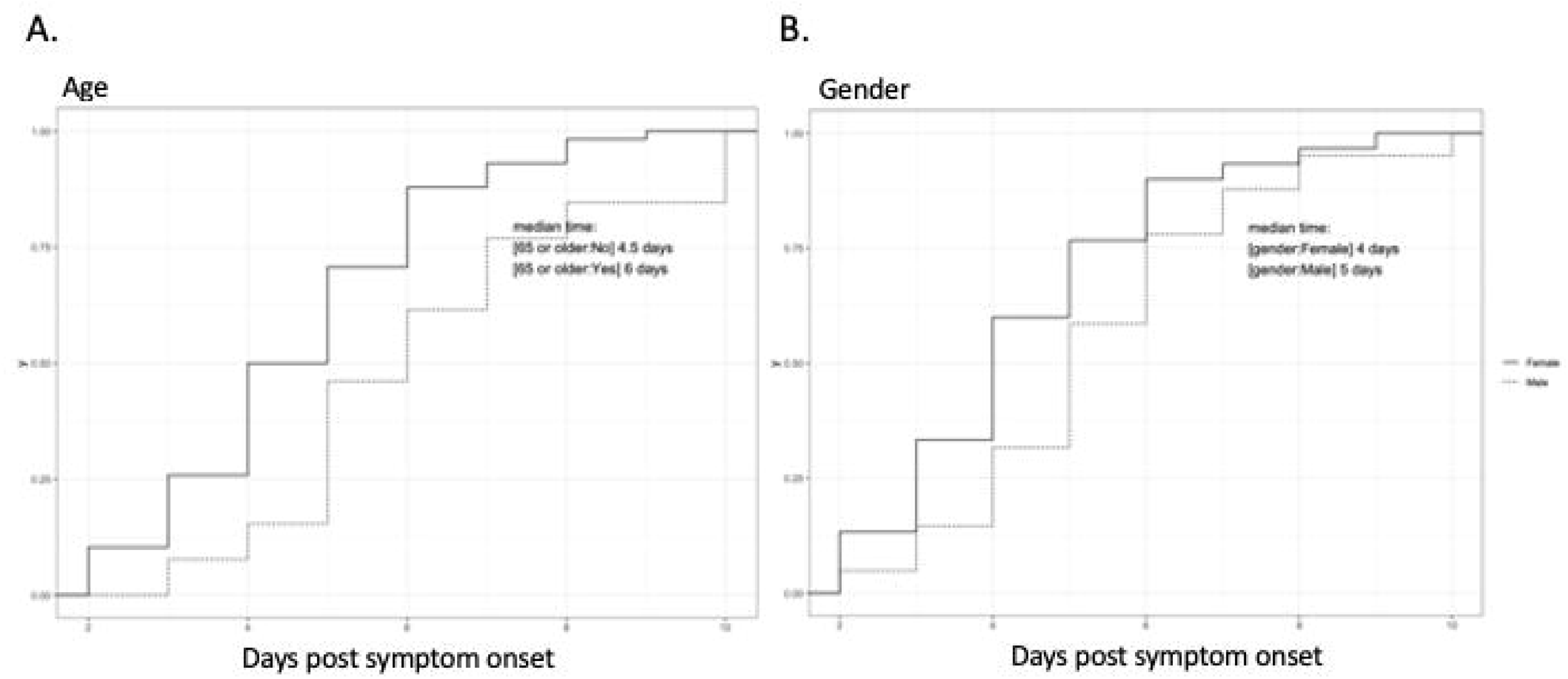
**A, B**. Association between the delay of triage visit from symptom onset and **A**) age and **B**) gender. The delay (days) is on the x-axis and the relative rank of the days is on the y-axis. A solid line was used for patients with age over 65 years and a dotted line for patients with age under 65 years. The solid line shows male patients and dotted line shows female patients.

